# Modelling testing frequencies required for early detection of a SARS-CoV-2 outbreak on a university campus

**DOI:** 10.1101/2020.06.01.20118885

**Authors:** Natasha K Martin, Robert T Schooley, Victor De Gruttola

## Abstract

**Background:** Early detection and risk mitigation efforts are essential for averting large outbreaks of SARS-CoV-2. Active surveillance for SARS-CoV-2 can aid in early detection of outbreaks, but the testing frequency required to identify an outbreak at its earliest stage is unknown. We assess what testing frequency is required to detect an outbreak before there are 10 detectable infections.

**Methods:** A dynamic compartmental transmission model of SARS-CoV-2 was developed to simulate spread among a university community. After introducing a single infection into a fully susceptible population, we calculate the probability of detecting at least one case on each succeeding day with various NAT testing frequencies (daily testing achieving 25%, 50%, 75%, and 100% of the population tested per month) assuming an 85% test sensitivity. A proportion of infected individuals (varied from 1–60%) are assumed to present to health services (HS) for symptomatic testing. We ascertain the expected number of detectable infections in the community when there is a > 90% probability of detecting at least 1 case. Sensitivity analyses examine impact of transmission rates (R_t = 0_ = 2, 2.5,3), presentation to HS (1%/5%/30%/60%), and pre-existing immunity (0%/10%)

**Results:** Assuming an 85% test sensitivity, identifying an outbreak with 90% probability when the expected number of detectable infections is 9 or fewer requires NAT testing of 100% of the population per month; this result holds for all transmission rates and all levels of presentation at health services we considered. If 1% of infected people present at HS and R_t=0_=3, testing 75%/50%/25% per month could identify an outbreak when the expected numbers of detectable infections are 12/17/30 respectively; these numbers decline to 9/11/12 if 30% of infected people present at HS. As proportion of infected individuals present at health services increases, the marginal impact of active surveillance is reduced. Higher transmission rates result in shorter time to detection but also rapidly escalating cases without intervention. Little differences were observed with 10% pre-existing immunity.

**Conclusions:** Widespread testing of 100% of the campus population every month is required to detect an outbreak when there are fewer than 9 detectable infections for the scenarios examined, but high presentation of symptomatic people at HS can compensate in part for lower levels of testing. Early detection is necessary, but not sufficient, to curtail disease outbreaks; the proposed testing rates would need to be accompanied by case isolation, contact tracing, quarantine, and other risk mitigation and social distancing interventions.

## BACKGROUND

SARS-CoV-2 continues to circulate in all states of the US and most foreign countries. In the absence of immunity in the vast majority of the population, even if the epidemic is effectively controlled over the coming summer, conditions are ripe for a return of the virus in the fall in winter.

Universities are struggling to determine whether and how to fully or partially reopen campus activities, particularly as universities and other high-density communities are at particular risk of explosive growth after viral introduction. Once transmission is established in a semi-closed university community with its dormitories, classrooms, student activities and multidisciplinary team-based research programs, growth of the epidemic would be expected to be rapid and control extremely difficult.

Widespread SARS-CoV-2 testing of active infection combined with case isolation of detected cases, comprehensive contact tracing and quarantine of contacts are critical tools for outbreak suppression. However, the level of NAT testing required to detect a SARS-CoV-2 outbreak at an early stage (below a threshold number of infections) is uncertain.

We conducted an epidemic modeling exercise to investigate the intensity of NAT testing required to achieve at least a 90% probability of detecting the presence of viral activity under different conditions for spread of virus from a single case within a university community. After detection of an outbreak, epidemiological measures including contact tracing and isolation of infected and exposed individuals might be able terminate the spread from this particular viral introduction within the community provided that the size of the outbreak is sufficiently small— but the conditions under which this could be achieved would require further investigation.

## METHODS

### Model

We developed a dynamic transmission model of SARS-CoV-2 infection and assessed the likelihood of detecting an infection on each successive day after the introduction of a single viral infection, given differing NAT testing volumes. The dynamic, deterministic, compartmental model was based on a susceptible-exposed-infected-recovered (SEIR) structure, with additional stratification of individuals by whether they were detectable and infectious (**Figure 1**, parameters and references in **Table 1**). Susceptible individuals (compartment **S**) can become infected through a dynamic process. Infected individuals were assumed to enter an exposed but not detectable or infectious stage (compartment **E**) for an average of 3 days. A proportion of individuals have disease that is either asymptomatic or sufficiently mild that they remain undiagnosed in the absence of active screening (compartment **A**); we assume they are infectious and detectable 3 days after infection and remain so for 14 days until recovery. The remaining proportion experience a short period of pre-symptomatic disease (compartment **P**), during which they are detectable and capable of transmission for 2 days prior to onset of symptoms. Upon developing symptoms (compartment **I**), individuals remain symptomatic and infectious for an average 2 days before presentation to health services (compartment **H**). Due to uncertainty regarding the proportion of students who are asymptomatic or mild enough disease such that they would not seek care, we explore several scenarios for this parameter in our analysis (1, 5%, 30%). Upon presentation to health services, we assume individuals are diagnosed and isolated, so no longer transmit infection. We assume that upon recovery individuals (compartment **R**), individuals were immune to further infection. We neglect mortality associated with COVID infection as this would not be expected to affect transmission dynamics at the earliest stage of the epidemic. The equations for the model are below:

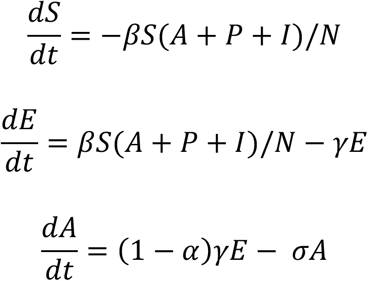

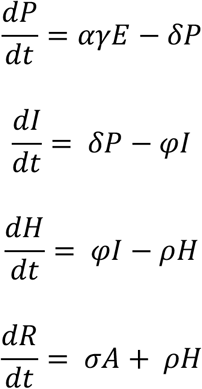

**Figure 1.**
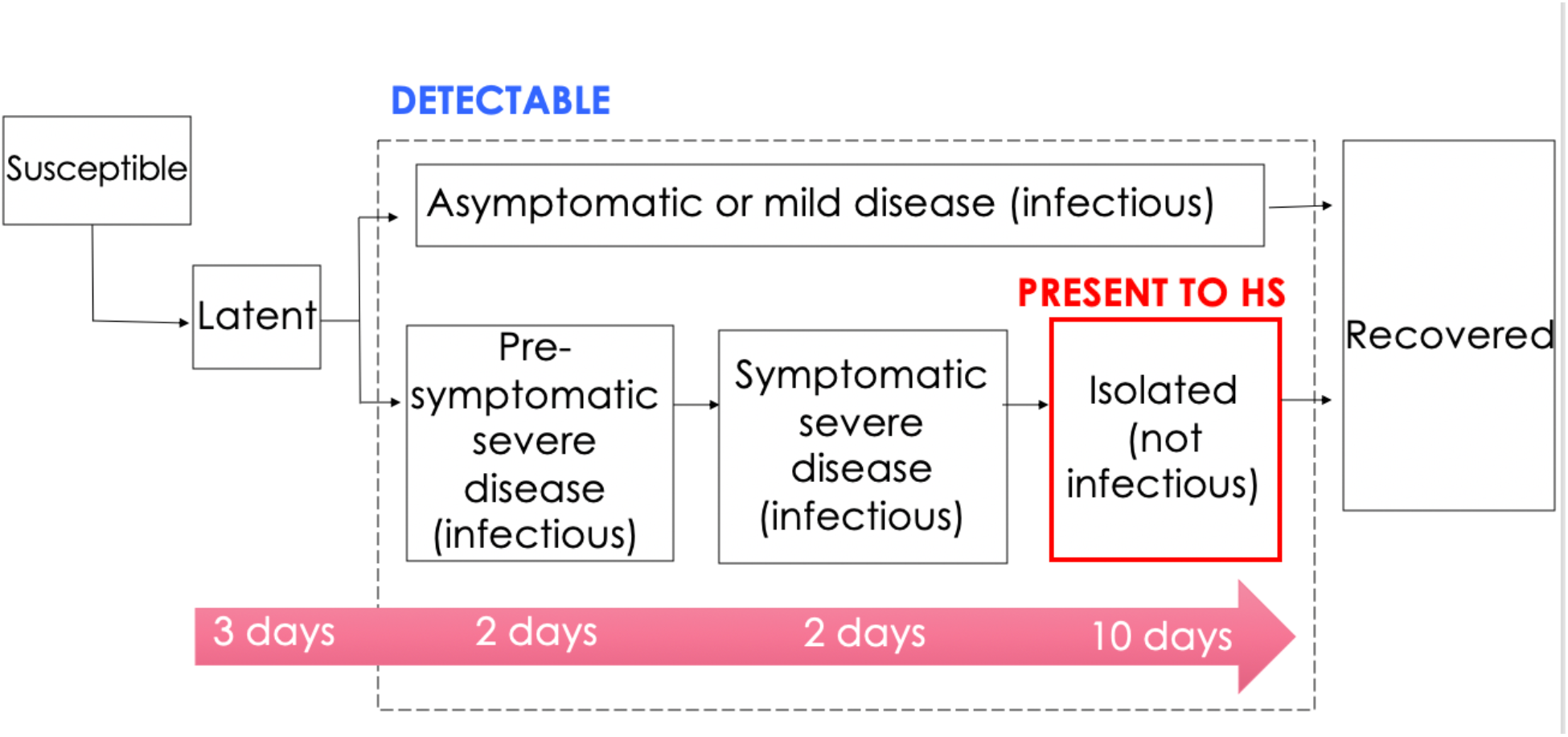
SARS-CoV-2 transmission model schematic. HS: health services.

**Table 1.** Model parameters.

| Parameter | Symbol | Value | Reference |
| --- | --- | --- | --- |
| Duration exposed but not detectable or infections | $1/\gamma$ | 3 days | 5 days incubation period[5], less 2 days viral shedding[6] |
| Duration detectable and infectious for asymptomatic or mild disease | $1/\sigma$ | 14 days | [6] |
| Duration pre-symptomatic but infectious and detectable for severe disease | $1/\delta$ | 2 days | [6] |
| Duration severe symptomatic until present to health services | $1/\varphi$ | 2 days | [7] |
| Duration infection after presentation to health services | $1/\rho$ | 10 days | Assumed 14 day total detection window[6] |
| Proportion who will eventually present to health services | $\alpha$ | 1%, 5%, 30%, 60% | Uncertain, varied |
| Transmission rate | $\beta$ | Varied to produce $R_{t=0} = 2, 2.5, 3$ | Uncertain and varies [8] |
| Test sensitivity | $s$ | 85% | [1] |
| Total population | $N$ | 65,000 | UCSD community |
| Daily number of tests | $d$ | % monthly tests * $N/30.5$ | Varied, 25%, 50%, 75%, 100% total population per month |

Where N = total population. The initial conditions are E(0) = 1, S(0) = 65000*(1-propImmune), R(0) = 65000*propImmune, A(0) = P(0) = I(0) = H(0) = 0. For our baseline analysis, propImmune = 0, but this is varied in the sensitivity analyses.

Using the epidemic model, we simulate outbreaks with the introduction of 1 infectious case to the university community assuming various transmission rates. The simulations predict: detectable cases (*A+P+I+H*), detectable cases not in contact with health services (*A+P+I*), and cumulative cases which have presented to health services for testing over time.

### Outbreak probability detection scenarios and calculations

We calculate the probability of detection of at least one case on each succeeding day after viral introduction with the following scenarios:

1. **HS:** Only passive testing upon presentation of symptomatic cases to health services
2. **25%/month**: 25% of the population tested per month (on an ongoing daily basis) plus passive testing upon presentation of symptomatic cases to health services
3. **50%/month**: 50% of the population tested per month (on an ongoing daily basis) plus passive testing upon presentation of symptomatic cases to health services
4. **75%/month**: 75% of the population tested per month (on an ongoing daily basis) plus passive testing upon presentation of symptomatic cases to health services
5. **100%/month**: Daily NAT testing resulting in 100% of the population tested per month plus passive testing upon presentation of symptomatic cases to health services

We calculate the probability of detection of at least one case on each succeeding day by assuming that the number of subjects who test positive is Poisson distributed—as is the number of symptomatic subjects who arrive at the clinic. The probability of detection of at least once case on each succeeding day, ***x***, is:

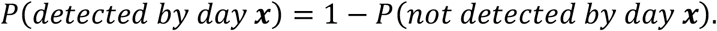

The probability of not detecting at least one case by each successive day, **x**, is the joint probability of not detecting an infection through presentation to HS by day **x**, and not detecting an infection through active testing by day **x**:

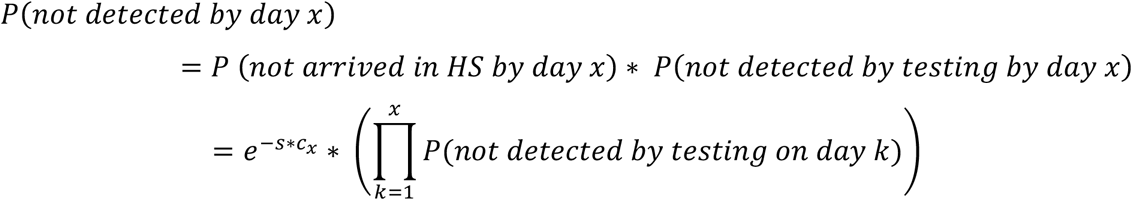

where c_x_ = expected number cumulative clinic arrivals by day **x**, and s = test sensitivity. The probability of not detecting an infection on each day by active testing is:

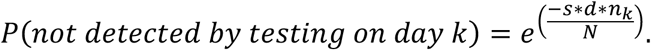

Where d = the number of daily tests, n_k_ = expected number detectable cases not in contact with health services on day **k**, and N = total population size. Hence, the overall probability of detection of at least one case on each successive day **x** is given by the expression:

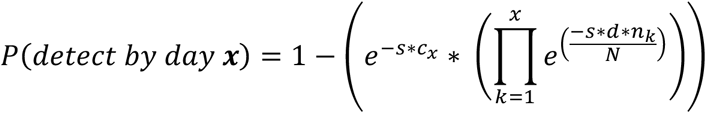

We vary the proportion of individuals who are symptomatic and seek care (1%, 5%, 30%,60%), and assume that the test sensitivity is 85% based on data from self-collected nasal or oral swabs[1]. We then determine the number of detectable infections present in the community on the day when there is a > 90% probability of detecting at least 1 case.

### Sensitivity analyses

We perform one-way sensitivity analyses to assess the impact of health services presentation rate (1%, 5%, 30%, 60%), transmission rate (R_t = 0_ = 2, 2.5,3), and pre-existing immunity (0%, 10%).

## RESULTS

The minimum number of detectable cases in the community when there is a > 90% probability of detecting at least 1 case is found in **Table 2** for various background health services (HS) presentation and transmission rates. With 100% testing of the community at a constant rate over each month and an 85% test sensitivity, the expected number of detectable cases in the community would be 6–9 (across scenarios) by the time we had at least 90% probability of detecting at least 1 case, regardless of background presentation rate to HS. If 75% of the community were tested each month, this expected number would be 7–12 detectable infections. If 50% of the community were tested each month, the expected number of detectable cases would be 9–15 if 5% present to HS, with more infections if 1% present to HS (10–17 detectable cases), and fewer if 30% or 60% present to HS (7–11 or 7–9 detectable cases, respectively). More infections were observed if only 25% of the community were tested each month (14–23 detectable infections for 5% presentation to HS).

**Table 2.** Minimum number of detectable cases in a community when there is a > 90% probability of detecting at least 1 case for various testing rates per month. Assumes 85% test sensitivity. *Combined with passive surveillance through presentation at health services. HS: health services.

|  | **Minimum number of detectable cases in a community when there is a >90% probability of detecting at least 1 case** |  |  |  |
| --- | --- | --- | --- | --- |
| **Percentage of UCSD community NAT tested per month\*** | **1% present to HS** | **5% present to HS** | **30% present to HS** | **60% present to HS** |
| **100%/month\*** |  |  |  |  |
| Low transmission ($R_{t=0}=2$) | 6 | 6 | 6 | 6 |
| Mid transmission ($R_{t=0}=2.5$) | 7 | 7 | 7 | 8 |
| High transmission ($R_{t=0}=3$) | 9 | 9 | 9 | 9 |
| **75%/month\*** |  |  |  |  |
| Low transmission ($R_{t=0}=2$) | 7 | 7 | 7 | 7 |
| Mid transmission ($R_{t=0}=2.5$) | 9 | 9 | 8 | 8 |
| High transmission ($R_{t=0}=3$) | 12 | 11 | 9 | 9 |
| **50%/month\*** |  |  |  |  |
| Low transmission ($R_{t=0}=2$) | 10 | 9 | 7 | 7 |
| Mid transmission ($R_{t=0}=2.5$) | 13 | 12 | 9 | 8 |
| High transmission ($R_{t=0}=3$) | 17 | 15 | 11 | 9 |
| **25%/month\*** |  |  |  |  |
| Low transmission ($R_{t=0}=2$) | 17 | 14 | 9 | 8 |
| Mid transmission ($R_{t=0}=2.5$) | 24 | 18 | 10 | 8 |
| High transmission ($R_{t=0}=3$) | 30 | 23 | 12 | 11 |
| **0%/month (only HS testing)** |  |  |  |  |
| Low transmission ($R_{t=0}=2$) | 149 | 34 | 10 | 8 |
| Mid transmission ($R_{t=0}=2.5$) | 198 | 45 | 13 | 9 |
| High transmission ($R_{t=0}=3$) | 229 | 54 | 14 | 11 |

Simulations showing the epidemic trajectory and day of detection are shown in Figure 2. The day of detection varied depending on transmission rate, with higher transmission rates resulting in shorter time to detection but also rapidly escalating numbers of cases in the absence of intervention. Higher background presentation rates to health services reduced the day of outbreak detection across all scenarios.

**Figure 2.**
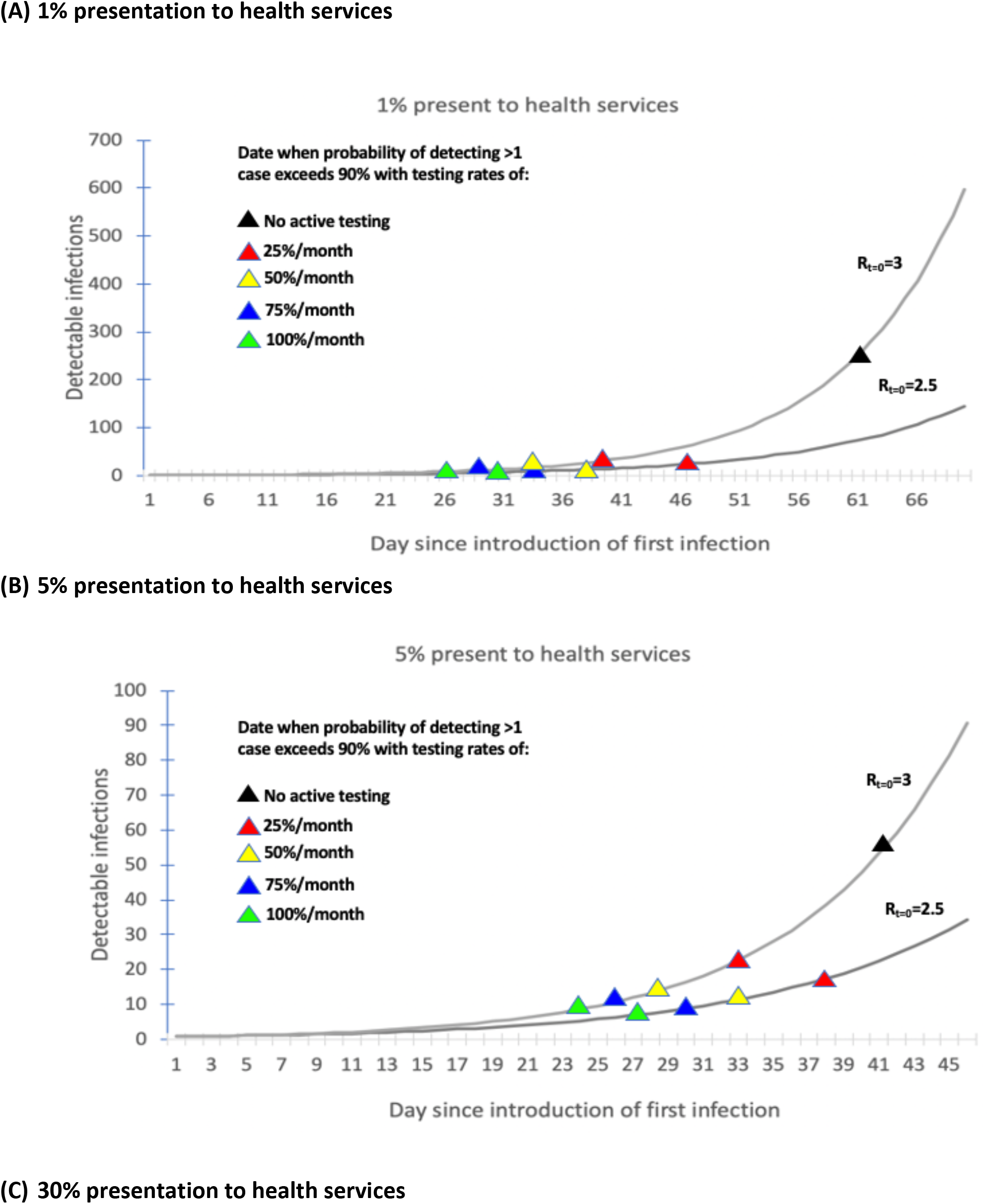
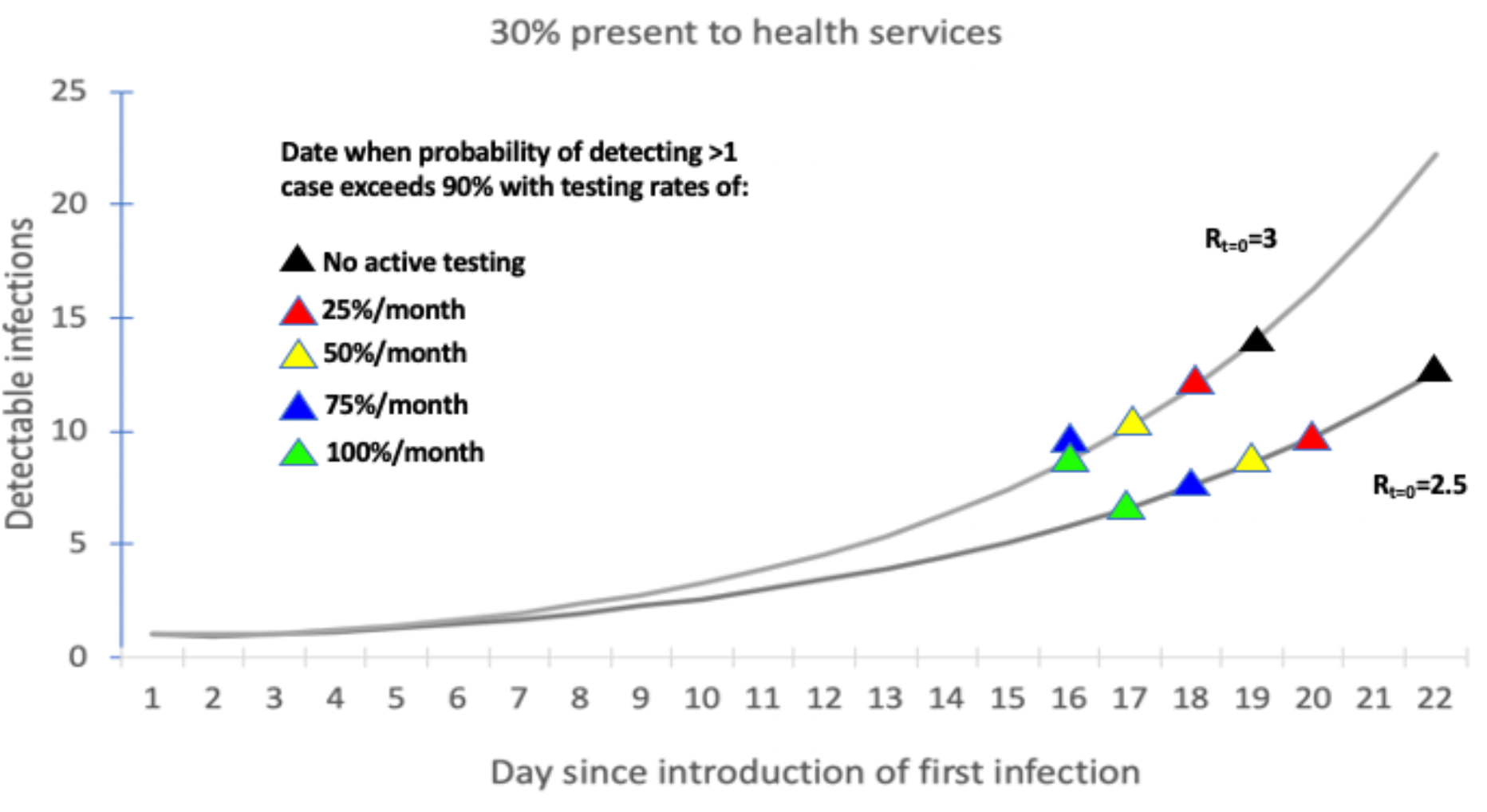
Effect of transmission rate on date of outbreak detection with various testing rates. HS: health services.

Active testing conferred important marginal benefits in the 1% and 5% presentation to HS scenarios (**Table 2, Figure 2a&b**). Without any active testing, the expected number of detectable cases in the community by the time we had at least 90% probability of detecting 1 case was the expected number of detectable cases was 149–229 (1% presentation to HS) and 34–54 (5% presentation to HS). Less marginal benefit was observed with 30% presentation to HS, where passive screening alone identified an outbreak with 10–14 detectable cases, 4–5 cases later than when combined with 100% active testing (**Table 2, Figure 2c**). Minimal marginal benefit was observed with 60% presentation to HS, where passive screening alone identified an outbreak with only 1–2 more cases than with 100% active testing (**Table 2**).

Higher levels of pre-existing immunity (**Table 3**) and higher test sensitivities (not shown) also reduced the number of detectable cases on the day of outbreak detection.

**Table 3.** Sensitivity analysis assuming a 10% immune at baseline. Assumes 85% test sensitivity. *Combined with passive surveillance through presentation at health services. HS: health services.

|  | **Minimum number of detectable cases in a community when there is a >90% probability of detecting at least 1 case** |  |  |  |
| --- | --- | --- | --- | --- |
| **Percentage of UCSD community NAT tested per month\*** | **1% present to HS** | **5% present to HS** | **30% present to HS** | **60% present to HS** |
| 100%/month* |  |  |  |  |
| Low transmission ($R_{t=0}=2$) | 6 | 6 | 6 | 6 |
| Mid transmission ($R_{t=0}=2.5$) | 7 | 7 | 7 | 8 |
| High transmission ($R_{t=0}=3$) | 9 | 9 | 9 | 9 |
| 75%/month* |  |  |  |  |
| Low transmission ($R_{t=0}=2$) | 7 | 7 | 7 | 7 |
| Mid transmission ($R_{t=0}=2.5$) | 9 | 9 | 8 | 8 |
| High transmission ($R_{t=0}=3$) | 12 | 11 | 9 | 9 |
| 50%/month* |  |  |  |  |
| Low transmission ($R_{t=0}=2$) | 10 | 9 | 7 | 7 |
| Mid transmission ($R_{t=0}=2.5$) | 13 | 12 | 9 | 8 |
| High transmission ($R_{t=0}=3$) | 17 | 15 | 11 | 9 |
| 25%/month* |  |  |  |  |
| Low transmission ($R_{t=0}=2$) | 17 | 14 | 9 | 8 |
| Mid transmission ($R_{t=0}=2.5$) | 24 | 18 | 10 | 9 |
| High transmission ($R_{t=0}=3$) | 30 | 23 | 12 | 11 |
| 0%/month (only HS testing) |  |  |  |  |
| Low transmission ($R_{t=0}=2$) | 149 | 34 | 10 | 8 |
| Mid transmission ($R_{t=0}=2.5$) | 198 | 45 | 13 | 9 |
| High transmission ($R_{t=0}=3$) | 228 | 54 | 14 | 11 |

## DISCUSSION

Early detection of viral outbreaks is essential to ensuring available resources for notification and isolation of cases, contact tracing, and quarantine. We find that monthly testing of 100% of the campus community is required to detect an outbreak when there are less than 9 or fewer detectable cases in the community, unless at least 30% of infected people develop symptoms that lead them to be tested HS. Even for the latter case, detection of outbreak when there are 9 or fewer detectable cases requires that 75% of the population be tested. As the percent tested declines, the size of the detectable populations increases fairly rapidly as testing levels decrease if the proportion of infected people is only 1 or 5%. But the increase is much slower if 30% or 60% do so; hence, monitoring of the university population for COVID-19 symptoms should be encouraged—especially if it is not feasible to implement high levels of testing.

Our study is the first to examine what NAT testing rates are required to detect a SARS-CoV-2 outbreak at an early stage. Other studies have examined what testing and isolation is required to prevent an outbreak or reduce the reproduction number to below 1, indicating that to prevent an outbreak with testing and case isolation alone testing of the entire population twice weekly is required[2]. Importantly, our analysis does not examine what is required to curtail the outbreak, only what is required to detect it at an early stage. At this point, epidemiological measures including contact tracing and isolation of infected and exposed individuals might be able terminate the spread of this particular viral introduction within the community—but investigation of the conditions under which this can be achieved is warranted. Previous mathematical models indicate that contact tracing and isolation of symptomatic contacts would likely require a very high rate of contacts traced in order to reduce the reproduction number to below 1, and could still result in very large outbreak sizes[3]. However, this same study indicates that contact tracing efficacy could be lower if initial outbreak sizes are smaller, supporting efforts to identify outbreaks at an earlier stage. Additionally, we did not examine the impact of isolation of asymptomatic contacts which is possible in a university setting and could provide additional risk mitigation benefit. Further, social distancing interventions could be implemented after the point of outbreak detection which could further reduce transmission[4].

### Limitations

#### Our study has a number of limitations

First, it is highly uncertain whether our proposed testing strategy is feasible and affordable. On a campus the size of UC San Diego, testing the entire campus community would require 65,000 individuals to be tested per month. We are currently implementing a campus testing program to assess feasibility of high-volume campus testing. This program is providing voluntary, free, NAT testing to resident undergraduate and graduate students on campus. Students drop into a testing site near to their residence hall and self-administer a nasal swab, which is collected by study staff and processed at an in-house CLIA-certified EUA compliant lab within 24–48 hours. Ongoing work is examining the logistics of expanding this program to the scale we propose. Saliva testing would likely increase willingness to participate. In addition to logistical scalability, cost is an important barrier. At the commercial price of $100 per test, this would equate to $6.5 million/month in testing alone. Our in-house lab charges substantially lower than commercial prices, but testing innovations are still needed to reduce this cost further.

Second, we assume that all individuals would be willing and available for testing. If those unwilling were not different in their patterns of transmissions than those who were willing, then we would expect the infection to fairly quickly cross over into our testing population. However, if the infection seeds within a highly connected network of individuals who all were unwilling to be tested then the outbreak size could be larger than we predict before detection.

Third, there is uncertainty in underlying parameters which we explore through sensitivity analyses, but still note that more robust data, particularly on the proportion accessing health services in the absence of active testing, would reduce uncertainty.

Fourth, we examine random testing strategies, but if high-risk sub-populations of students could be ascertained, then targeted testing strategies could be developed which could improve testing efficiency (requiring fewer tests to identify outbreaks). A key question is what subgroups are higher-risk, and whether and how these risks are associated with clusters in the contact network. For example, do students coming from high-risk areas tend to share more classes together? Collection of routine testing data combined with information on risk and contact networks would allow us to better target testing, possibly through an adaptive approach as these networks change over time.

### Conclusion

Early SARS-CoV-2 outbreak detection is achievable with high rates of NAT testing (100%/month) or possibly with lower rates of testing but a high proportion of infected people—even those with mild symptoms-- presenting at HS for testing. Testing needs to be combined with intensive case isolation, contact tracing and quarantine, and social distancing recommendations to curtail the observed outbreak and reduce risk of further transmission.

## Data Availability

All data available on request

## Funding Acknowledgements

This study was funded by a University of California COVID-19 Emergency Seed Grant. NKM additionally acknowledges support from NIAID and NIDA (R01AI147490) and the University of San Diego Center for AIDS Research (CFAR), a NIH funded program (P30 AI036214). RTS is supported by NIAID (R01 AI131424).

